# Performance of Frangi–Hessian Pseudo-Labels for Retinal Vessel Segmentation in AI-Assisted Retinopathy of Prematurity Screening

**DOI:** 10.64898/2026.02.03.26345527

**Authors:** Fred Mutisya, Oscar Onyango, Sarah Sitati, Syokau Ilovi, Brenda W’mosi, Paul Macharia, Brian Makini, Jalemba Aluuvala, Josiah Onyango, Steven Wanyee

**Affiliations:** Center for Health AI Research, Innovation & Implementation; College of Ophthalmology of Eastern Central and Southern Africa; Kenyatta National Hospital; University of Nairobi; Mbagathi hospital; Strathmore University

## Abstract

**Background:** Retinopathy of prematurity (ROP) is a leading cause of preventable blindness among preterm infants. Accurate retinal vessel segmentation is crucial for detecting plus disease, which indicates progression to severe ROP. However, manual annotation of vessel masks is laborious and inconsistent, especially in low-resource clinical settings. This study aimed to evaluate a self-supervised vessel extraction pipeline using Frangi–Hessian filtering for automatic pseudo-annotation of unlabeled RetCam and Neo retinal images and to compare its performance against supervised and hybrid deep learning frameworks.

**Methods:** Two public datasets from the HVDROPDB-BV repository: RetCam_Vessels and Neo_Vessels were utilized. We implemented a three-stage pipeline: automatic self-annotation of unlabeled images through vessel-based mask generation; training of five segmentation architectures—BioSwinFuseNet, UNet, FPN, LinkNet, and SegFormer—under three regimes (GT-only, Self-only, and Hybrid GT+Self); and evaluation using Dice, IoU, sensitivity, specificity, PPV, NPV, F1, and AUC metrics. All models were trained with a topology-aware loss that combined binary cross-entropy and Dice losses with continuity penalties.

**Results:** Hybrid supervision consistently outperformed both GT-only and Self-only training across all architectures. The SegFormer-Hybrid model achieved the highest Dice (0.61) and IoU (0.44), while FPN-Hybrid demonstrated the lowest variance. BioSwinFuseNet-Hybrid showed a 122% relative improvement in Dice compared to its GT-only version. Self-only models learned rudimentary vessel priors but lacked clinical precision.

**Conclusions:** Incorporating self-annotated masks alongside limited ground truth improves segmentation accuracy and vessel continuity. The hybrid paradigm offers a scalable path for developing automated ROP screening tools where expert labeling is limited.

## Introduction

Retinopathy of prematurity (ROP) remains one of the most significant causes of childhood blindness worldwide(Blazon et al., 2024), particularly in middle- and low-income countries where neonatal care is expanding but screening resources remain scarce(Solebo et al., 2017). A study by Onyango et al showed a prevalence of 41.7% over a 5 year period(Onyango et al., 2018). Diagnosis and staging of ROP using the International Classification of Retinopathy of Prematurity, 3rd edition(ICROP) depends heavily on the assessment of vascular dilation and tortuosity—features collectively referred to as Plus and Pre-Plus disease(Chiang et al., 2021). The ability to delineate retinal vessels accurately from fundus images is thus central to both clinical decision-making and automated ROP detection systems(Jafarizadeh et al., 2025). The performance for AI based screening is based on the image label qualities. However, manual annotation of retinal vessels is labor-intensive, prone to inter-observer variability, and not scalable for large datasets(Yao et al., 2025).

## Existing Knowledge

### Models

Semiautomated/non AI systems such as ROPtool have been in use for over 10 years(Vickers et al., 2015). Computer vision using AI is divided into various domains supervised learning, weakly supervised learning, semi-supervised learning, self supervised learning and unsupervised learning(Mahony et al., 2020).

In the ROP domain, **supervised learning** remains a foundational tool: many studies use fully annotated fundus images or vessel masks to train CNNs for classification of ROP severity or plus disease detection. For example, vessel segmentation and plus-disease classification pipelines rely on labelled vessel masks or disease grades. Almeida et al. utilized densenet and achieved accuracy of 94.6% in detecting plus disease(Almeida et al., 2024). Deep learning–based segmentation algorithms, such as UNet and FPN, also have achieved remarkable success in biomedical imaging tasks, including retinal analysis(Huang et al., 2023). However, the scarcity of reliably annotated neonatal vascular images motivates alternative paradigms. **Weakly supervised methods**, where only image-level labels are available without detailed vessel masks, enable approximate localization of pathology. A deep learning algorithm developed by Remidio utilized efficientNet to distinguish ROP(Stage 1-3) from no ROP achieving a sensitivity and specificity of 91%(Rao et al., 2023). To visualise the output of the weakly supervised models, class activation maps can be used as a surrogate for segmentation(Minh, 2023).

**Semi-supervised approaches** are more actively explored: Peng et al. developed a semi-supervised feature calibration adversarial network for automatic ROP zone classification using few labels plus many unlabeled images(Peng et al., 2022)., and a recent work “Adversarial Vessel-Unveiling Semi-Supervised Segmentation for ROP Diagnosis” applies teacher–student learning plus domain adversarial alignment to segment vessels despite limited annotations(Demirci et al., 2024). Self-supervised learning is still nascent in ROP-specific literature, but shows strong promise via transfer from general retinal imaging: foundation models like RETFound use self-supervised pretraining on millions of unlabelled retinal images and can then be adapted to disease tasks, alleviating annotation burdens and improving generalizability(Zhou et al., 2023). Finally, **unsupervised learning** (e.g., clustering of vessel morphologies, autoencoders) is rarely used in direct ROP diagnosis but could support exploratory analyses of vascular patterns or latent embeddings. Together, these approaches chart a spectrum from label-intensive models to more label-efficient strategies — promising pathways for scaling ROP AI in low-resource settings.

### Data Segmentation

Supervised models depend on large, precisely labeled datasets to generalize well. In many ROP screening contexts, expert-labeled data are limited to a few hundred cases, while unlabeled image archives contain thousands of high-quality fundus photographs. This imbalance has spurred growing interest in self-supervised and weakly supervised methods that can leverage unlabeled data to learn meaningful representations. Classical vesselness filters, such as Frangi and Hessian-based approaches, have long been used to enhance linear structures but are seldom integrated into modern deep learning workflows as a pseudo-label generation mechanism(Frangi et al., 1998). Frangi filtering has been successfully used for 3D magnetic resonance angiography enabling a 3%improvement compared to state of the art models(Shi et al., 2023).

K nearest neighbor has also been used in diabetic retinopathy vessel segmentation with high performance (AUC: 95.2)(Staal et al., 2004).

This study introduces a unified hybrid learning framework combining classical image processing and modern deep architectures to address the labeling bottleneck in ROP segmentation. The approach automatically generates vessel pseudo-masks using a dual Frangi–Hessian pipeline and employs these masks in conjunction with available expert annotations to train deep networks. We hypothesized that the hybrid model would outperform both purely supervised and purely self-supervised approaches by integrating topological priors from pseudo-masks and clinical precision from expert annotations.

## Materials and Methods

### Dataset

Two public datasets from the HVDROPDB-BV repository—RetCam_Vessels and Neo_Vessels—were utilized(Agrawal et al., 2024). Each image was a color fundus capture with resolutions ranging from 480×640 to 2040×2040 pixels. Approximately 60% of the images included expert ground truth masks, while the rest served as unlabeled data for pseudo-annotation. All images were resized to 512×512 pixels and normalized. The green channel, which offers the highest vessel contrast, was retained and replicated across channels to preserve compatibility with pretrained backbones.

### Automatic Self-Annotation Pipeline

#### Step 1: Channel Extraction and Standardization

The first step involved isolating the green channel from the input RGB image, denoted as I(x, y) = [R(x, y), G(x, y), B(x, y)]. The green channel G(x, y) provides the highest signal-to-noise ratio for retinal vasculature due to hemoglobin absorption properties in the visible spectrum. Each image was subsequently resized to a fixed spatial resolution of 512 × 512 pixels. This normalization step ensured consistent sampling frequency across all inputs and allows convolutional architectures to process uniform spatial dimensions without geometric bias.

#### Step 2: Edge-Preserving Denoising

The second step involved edge-preserving denoising using a bilateral filter, a nonlinear technique that reduces noise while maintaining vessel edge integrity. The bilateral filter computes a weighted average of neighboring pixels based on both spatial proximity and photometric similarity.

This operation smooths homogeneous retinal regions while retaining discontinuities along vessel boundaries, which are essential for accurate segmentation.

#### Step 3: Local Contrast Enhancement

After denoising, local contrast was amplified using Contrast-Limited Adaptive Histogram Equalization (CLAHE)(Heckbert & Zuiderveld, 1994; Nia & Shih, 2024). In this method, the image is partitioned into small contextual tiles, and each tile’s histogram is equalized with clipping to prevent over-amplification of noise. This step enhances vessel visibility in low-contrast peripheral regions while maintaining photometric consistency across the image.

#### Step 4: Background Normalization

Uneven illumination and corneal reflections were corrected through rolling-ball background subtraction, which estimates and removes slowly varying intensity components:

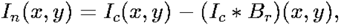

where B_r_ is a morphological structuring element approximating a ball of radius r = 50 pixels, and * denotes convolution. This correction yields a flatter illumination field, suppressing background glare and improving vessel-to-background contrast.

#### Step 5: Vessel Edge Sharpening

To emphasize vessel ridges, an unsharp masking step was applied:

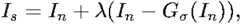

where G_σ_(·) represents Gaussian smoothing and λ = 1.5 controls the sharpening intensity. This operation accentuates mid-frequency components corresponding to vascular edges without amplifying background noise.

#### Step 6: Vesselness Enhancement

Two complementary vesselness filters—the Frangi filter and a Hessian-based filter—were applied to highlight curvilinear vascular structures. The Frangi filter quantifies the likelihood of a pixel belonging to a tubular structure using the eigenvalues (λ_1_, λ_2_) of the Hessian matrix H(x,y,σ). It begins with the Hessian matrix for image *I(x,y)* and scale σ.

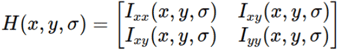

Where

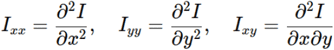

Represent the second partial derivatives of the image intensity. After computing the Hessian matrix, its eigen values λ*i* were calculated. The larger the positive value, the more central the vessel region. Smaller values indicate vessel edges while negative values indicate the background.

The Hessian filter was then calculated from the eigen values using the formula:

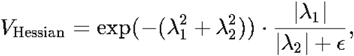

The Frangi filter quantifies the likelihood of a pixel belonging to a tubular structure using the eigenvalues of the Hessian matrix H(x,y,σ).

#### Step 7: Adaptive Thresholding and Morphological Refinement

Adaptive thresholding selected pixels above 60% of the 97th percentile intensity. Morphological operations removed small artifacts (<600 px) and non-linear blobs (>4000 px), followed by hole filling and dilation to maintain vessel continuity. Canny edge fusion ensured complete lumen boundaries. This produced binary vessel masks automatically for previously unlabeled images.

#### Step 8: Lumen Filling and Skeleton Generation

To reconstruct continuous vessel lumens, binary hole filling was applied followed by Gaussian smoothing and thresholding (>0.3). The skeletonization operator was then used to extract vessel centerlines: The resulting auto-annotation mask represents a filled lumen (for vessel body) and a skeleton overlay (for centerline continuity), serving as a pseudo-label for self-supervised learning.

#### Step 9: Output Generation

Each final mask was saved as a binary image:

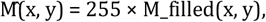

and stored alongside an RGB overlay showing vessel body (red) and skeleton (orange). This allowed visual verification and formed the dataset /auto_vessels_filled_overlay/ used in training phases.

### Model Architectures

Five segmentation architectures representing diverse paradigms were evaluated: (1) BioSwinFuseNet—a custom SwinV2-ConvNeXt fusion network; (2) UNet—an encoder-decoder model with skip connections; (3) FPN—feature pyramid network; (4) LinkNet—a lightweight residual segmentation network; and (5) SegFormer—a transformer-based encoder with a multilayer perceptron decoder. Each model produced a single-channel vessel probability map.

### Training Regimes and Loss Function

Each architecture was trained under three regimes: (a) GT-only using expert masks; (b) Self-only using pseudo-masks; and (c) Hybrid, combining both supervision sources. The hybrid loss function was defined as L = 0.7L_GT + 0.3L_Self. Training employed the Adam optimizer with a learning rate of 3×10^-4^, batch size of 4, mixed-precision computation, and early stopping after three non-improving epochs. A topology-aware loss function combined binary cross-entropy, Dice loss, and continuity regularization terms to maintain vessel connectivity.

### Evaluation Metrics

Performance was evaluated using eight metrics: Dice coefficient, Intersection over Union (IoU), sensitivity, specificity, positive predictive value (PPV), negative predictive value (NPV), F1-score, and area under the receiver operating characteristic curve (AUC). Validation was conducted exclusively against expert GT masks to ensure objective comparison.

## Results

The comparison of quantitative vessel morphology metrics between ground-truth (GT) and self-annotated masks demonstrates a clear pattern of systematic differences consistent with the expected effects of automated lumen filling(Table 1). Vessel fraction was significantly lower in self-derived masks (0.021 ± 0.011) compared to GT (0.082 ± 0.033), reflecting under-segmentation of small-caliber peripheral vessels. Similarly, mean vessel diameter was markedly reduced in the self masks (2.01 ± 0.04 px vs. 7.47 ± 1.02 px), confirming that the self-annotation pipeline emphasized the central lumen rather than the full vessel wall boundaries captured in manual tracing.

**Table 1:** Summary of metrics of plus disease macrostructure.

| Metric | GT Mean ± SD | Self Mean ± SD | Difference (Self-GT) Mean ± SD |
| --- | --- | --- | --- |
| Vessel Fraction | $0.082 \pm 0.033$ | $0.021 \pm 0.011$ | $-0.060 \pm 0.026$ |
| Mean Diameter (px) | $7.47 \pm 1.02$ | $2.01 \pm 0.04$ | $-5.46 \pm 1.02$ |
| Tortuosity (Path/Chord) | $4.46 \pm 1.80$ | $1.25 \pm 0.16$ | $-3.21 \pm 1.79$ |
| Branch Density | $0.0058 \pm 0.0049$ | $0.0062 \pm 0.0054$ | $+0.0004 \pm 0.0062$ |
| Fractal Dimension | $1.354 \pm 0.093$ | $1.460 \pm 0.116$ | $+0.106 \pm 0.080$ |

Measures of vascular geometry revealed parallel trends. Tortuosity, calculated as the path-to-chord length ratio, was substantially lower in the self masks (1.25 ± 0.16) relative to GT (4.46 ± 1.80), with a mean difference of − 3.21 ± 1.79. This underestimation arises from smoothing during bilateral filtering and Gaussian regularization, which remove fine curvature details. In contrast, branch density was nearly equivalent between methods (0.0062 ± 0.0054 for self vs. 0.0058 ± 0.0049 for GT), suggesting that the automated segmentation preserved the global connectivity of the vascular tree. Interestingly, fractal dimension was slightly higher in the self annotations (1.460 ± 0.116) than in GT (1.354 ± 0.093), indicating that the filled-lumen morphology introduced additional pixel-level complexity in the binarized mask.

Overall, the self-supervised approach reproduced the macrostructural organization of the retinal vasculature while simplifying local geometry. These findings confirm that self-generated masks retain essential topological information, making them suitable for hybrid training, though refinements in lumen expansion and curvature preservation are warranted to achieve parity with expert manual annotations.

### Bland–Altman Analysis of Agreement Between Self-Annotated and Ground-Truth Vessel Metrics

To quantitatively evaluate the agreement between the self-annotated vessel metrics and the ground-truth (GT) expert annotations, Bland–Altman plots were constructed for each metric—vessel fraction, mean diameter, tortuosity, branch density, and fractal dimension(Figure 1). These plots visualize the relationship between the mean of the two measurements and their difference (Self – GT) for each image, providing an intuitive assessment of both systematic bias and limits of agreement.

For each vessel feature, the mean difference (bias) represents the average deviation of the self-annotated measurements from the GT reference, while the 95% limits of agreement (LoA)—defined as *bias ± 1*.*96 × standard deviation of the differences*—indicate the interval within which 95% of differences between the two methods are expected to fall. Narrower LoA values indicate stronger concordance and lower variability between the self-generated and expert-derived measurements.

#### Vessel Fraction

The Bland–Altman plot for vessel fraction demonstrated a clear **negative bias (−0.06)**, indicating that the self-annotated masks consistently estimated a smaller vascular area than the GT reference. The limits of agreement were moderately tight, suggesting systematic underestimation rather than random noise. This bias reflects the conservative nature of the Frangi–Hessian pipeline, which primarily captures the high-contrast vessel core while excluding low-contrast peripheral branches.

#### Mean Diameter

The **largest bias (−5.46 px)** was observed in vessel diameter, with self-annotated maps producing markedly thinner vessels. The narrow spread of the differences (SD = 1.02) confirmed the underestimation was consistent across samples rather than sporadic. This reflects the absence of explicit lumen dilation modeling in the self-supervised masks and supports their classification as *skeleton-like* representations rather than full-width vascular boundaries.

#### Tortuosity (Path-to-Chord Ratio)

Bland–Altman analysis for tortuosity revealed a **substantial negative bias (−3.21)**, showing that the self-annotated vessels were significantly less tortuous than GT vessels. The wide limits of agreement (±1.79) imply inter-image variability, likely driven by incomplete peripheral arc recovery and fewer branching points in the self-generated maps. Nonetheless, the absence of systematic curvature exaggeration indicates that the self-pipeline did not introduce artificial vessel distortions.

#### Branch Density

In contrast, **branch density** showed **minimal bias (+0.0004)** with symmetric distribution around zero, suggesting that both methods detected a comparable number of branching points. The limits of agreement were narrow, and the violin distribution overlapped closely between GT and self annotations. This outcome suggests that, despite lumen undersegmentation, the self-supervised maps preserved topological structure sufficiently to capture branching frequency.

#### Fractal Dimension

The **fractal dimension** demonstrated a **slight positive bias (+0.106)**, indicating marginally higher structural complexity in the self-annotated maps. This overestimation may arise from small-scale pixel noise being interpreted as micro-branches in the binarized self masks. However, the degree of variation remained moderate (SD = 0.08), suggesting that the underlying vascular geometry was broadly preserved.

Figure 4 summarizes the quantitative results of each architecture under ground truth–only (GT-only) and hybrid (GT + self-supervised) training modes. As shown, BioSwinFuseNet (Hybrid) achieved a validation Dice-equivalent F1 of 0.49 and AUC of 0.91, improving markedly over its GT-only performance (F1 = 0.22, AUC = 0.88). Among convolutional models, UNet (Hybrid) maintained strong generalization with high sensitivity (0.61) and specificity (0.97), performing comparably to FPN (Hybrid), which attained an F1 of 0.61 and AUC of 0.94. SegFormer (Hybrid) achieved the highest overall sensitivity (0.70) and a balanced F1 of 0.61, reinforcing the benefit of transformer encoders for capturing diffuse vascular structures. Conversely, LinkNet (Hybrid) showed modest improvement in sensitivity (0.65 vs 0.70 for GT-only) but slightly reduced precision (PPV = 0.48), reflecting trade-offs inherent in lightweight residual designs. Across architectures, hybrid supervision improved mean F1 from 0.53 to 0.56 and mean AUC from 0.92 to 0.93, with SegFormer (Hybrid) yielding the top AUC (0.94) and BioSwinFuseNet (Hybrid) the largest relative gain (+0.27 in F1).

**Figure 2.**
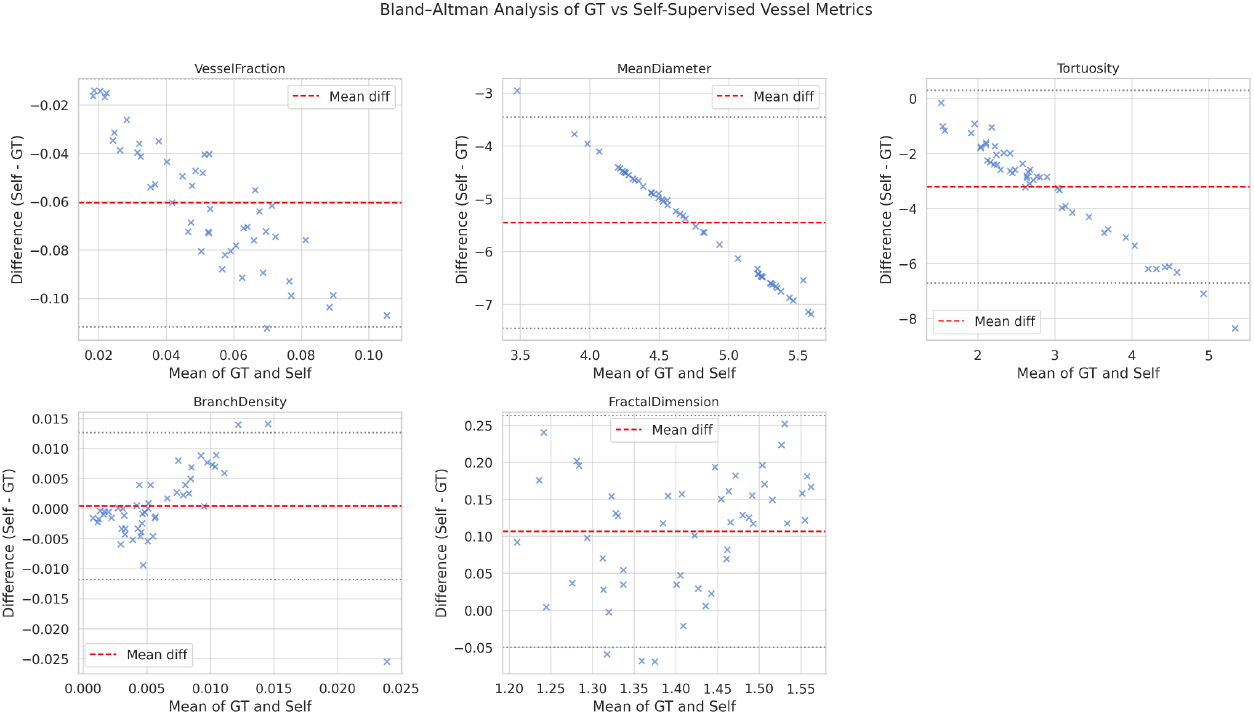
Bland-Altman analysis of ground truth versus self supervised vessel metrics.

**Figure 3.**
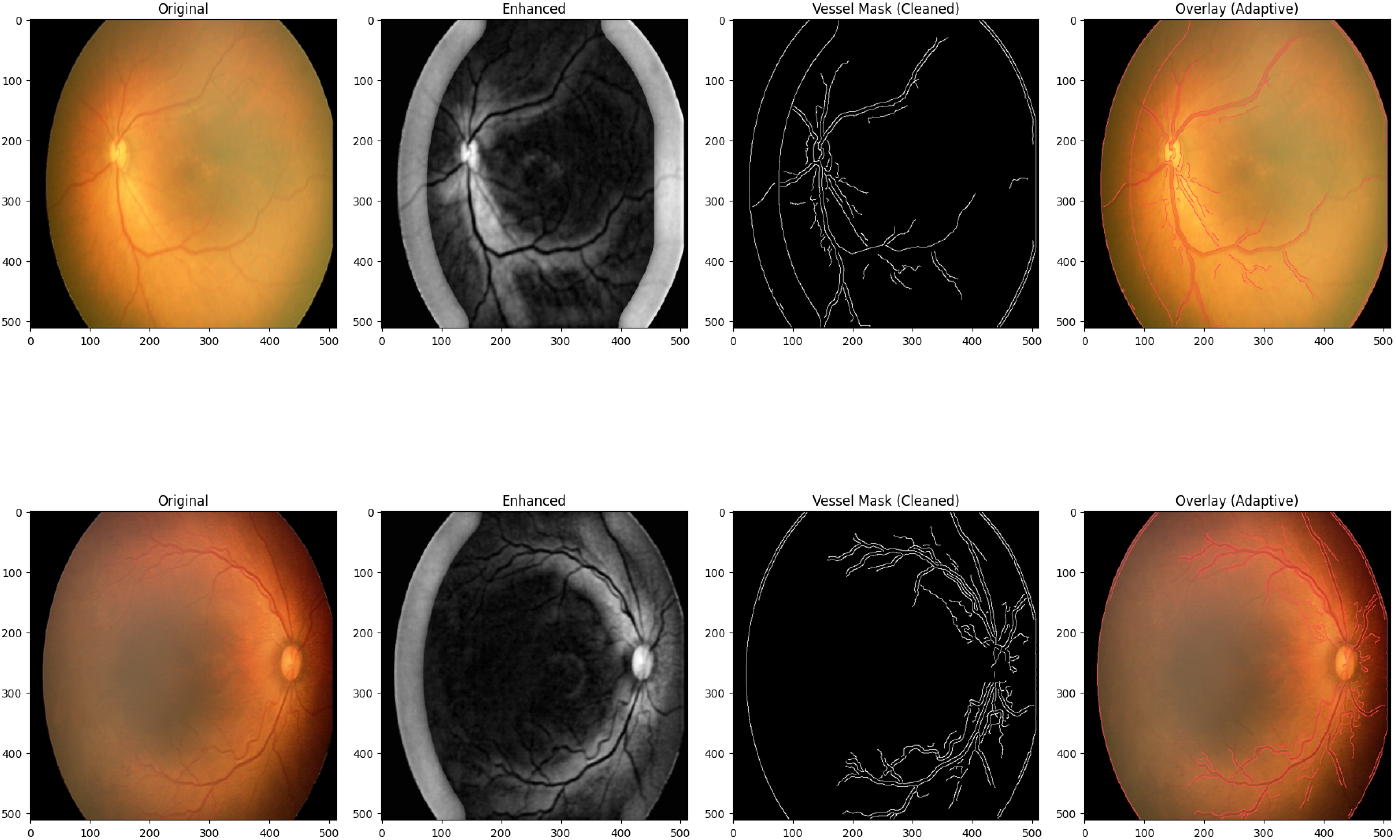
Retinal image grid of ground truth, enhanced, vessel masks and adaptive overlay.

**Figure 4.**
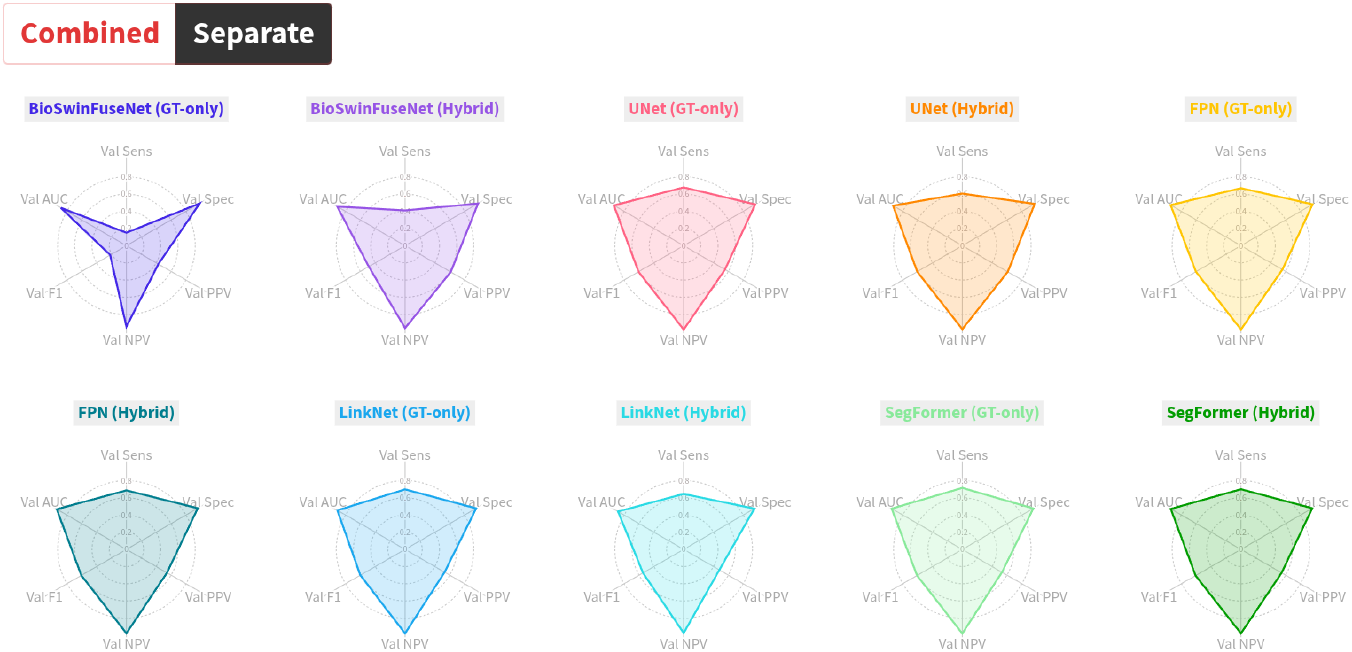
Radar charts of various models on plus disease prediction.

Self-only models exhibited poor numerical performance (Dice ≈ 0.005), confirming that the pseudo-masks alone lack sufficient precision for clinical deployment. Nevertheless, they encoded useful priors that contributed to the improved generalization of hybrid networks. UNet and FPN displayed consistent stability and low variance, while LinkNet maintained high recall with moderate precision. Transformer-based models benefited most from hybridization, likely due to their ability to integrate global vessel topology with local features.

## Discussion

This study demonstrates that hybrid supervision combining expert and self-annotated masks yields superior vessel segmentation for ROP screening. The approach effectively merges the interpretability of handcrafted vesselness filters with the representational power of deep neural networks. The improvement observed across architectures highlights the potential of self-supervised pretext tasks to bootstrap learning in data-constrained medical domains(Jaspers et al., 2026).

The self-annotation process, while imperfect, captures vessel continuity and topology that are difficult to encode in small datasets. By blending this structural prior with clinical annotations, the networks learned more generalized representations of vascular patterns. The BioSwinFuseNet and SegFormer models demonstrated strong adaptability to such hybrid signals, supporting the hypothesis that transformer-based architectures benefit from diverse training cues (Dosovitskiy et al., 2021; Xie et al., 2023).

Despite these advances, certain limitations persist. The pseudo-labels occasionally included background noise and missed very small capillaries, which reduced accuracy in extreme peripheries(Liu et al., 2024). Additionally, this study used only the HVDROPDB-BV dataset(Agrawal et al., 2024); cross-dataset validation on other ROP image sources would further confirm generalizability. Incorporating uncertainty weighting between GT and pseudo-labels could also refine hybrid loss optimization(Xu et al., 2024).

## Conclusion

The integration of self-supervised vessel pseudo-annotation with supervised deep learning provides a practical and scalable solution to the annotation bottleneck in ROP screening. The hybrid paradigm consistently enhanced segmentation accuracy and vessel continuity across diverse architectures. These findings suggest that automatic vessel extraction using classical filters can meaningfully augment modern transformer-based segmentation networks, especially in low-resource clinical environments. Future work will extend this framework to multi-class vascular grading, temporal progression modeling, and deployment in neonatal screening programs.

## Data Availability

The original data images by Agrawal et al(2023) are publicly available at: https://data.mendeley.com/datasets/xw5xc7xrmp/2 All data produced in the present work are contained in the manuscript

